# Awake prone positioning effectiveness in moderate to severe COVID-19 a randomized controlled trial

**DOI:** 10.1101/2024.06.30.24309722

**Authors:** Nguyen Thanh Phong, Du Hong Duc, Ho Bich Hai, Nguyen Nguyen Thanh, Le Dinh Van Khoa, Le Thuy Thuy Khanh, Luu Hoai Bao Tran, Nguyen Thi My Linh, Cao Thi Cam Van, Dang Phuong Thao, Nguyen Thi Diem Trinh, Pham Tieu Kieu, Nguyen Thanh Truong, Vo Tan Hoang, Nguyen Thanh Ngoc, Tran Thi Dong Vien, Vo Trieu Ly, Tran Dang Khoa, Abigail Beane, James Anibal, Oucru Covid Reseach Group, Guy E Thwaites, Ronald B Geskus, David Clifton, Nguyen Thi Phuong Dung, Evelyne Kestelyn, Guy Glover, Le Van Tan, Lam Minh Yen, Nguyen Le Nhu Tung, Nguyen Thanh Dung, C Louise Thwaites

## Abstract

**Objectives:** We evaluated the efficacy and acceptability of awake-prone positioning (APP) in a randomised controlled trial, using a dedicated APP implementation team and wearable continuous-monitoring devices to monitor position and oximetry.

**Methods:** The trial was performed at a tertiary level hospital in Ho Chi Minh City, Vietnam, recruiting adults (≥18 years) hospitalised with moderate or severe COVID-19 and receiving supplemental oxygen therapy via nasal/facemask systems or high-flow nasal canulae. Participants were randomized (1:1) to standard care or APP. The primary outcome was escalation of respiratory support within 28 days of randomisation.

**Results:** Ninety-three patients were enrolled between March 2022 and March 2023; 80 (86%) had received ≥2 doses of SARS-CoV2 vaccine. Significantly greater mean daily APP times were achieved in those allocated to APP, although most did not achieve the target 8 hours/day. We did not detect significant differences in the primary outcome (RR 0.85, 95% CI 0.40-1.78, p=0.67) or secondary outcomes, including intubation rate and 28-day mortality. Particpants reported prone positioning was comfortable, although almost all preferred supine positioning. No adverse events associated with the intervention were reported.

**Conclusions:** APP was not associated with benefit, but was safe. Continuous monitoring with wearable devices was feasible and acceptable to patients.

**Clinical Trials Registration:** NCT05083130

**Funding:** Wellcome Trust Grant 089276/B/09/7, 217650/Z/19/Z and FDCO/Wellcome Trust 225437/Z/22/Z

## Introduction

In mechanically ventilated patients with acute respiratory distress syndrome (ARDS), prone positioning is associated with improved survival (1). Benefit of prone positioning in patients not receiving invasive mechanical ventilation is less clear, but the COVID-19 pandemic stimulated multiple randomised controlled trials of prone position in non-mechanically ventilated patients, termed awake prone positioning (APP). Comparison of these studies is enabled by consistent use of similar endpoints and several studies using a harmonised protocol (2). Nevertheless, results have shown conflicting results. Whilst a meta-analysis showed overall benefit of APP in patients with COVID-19, subgroup analysis showed no benefit in those treated outside ICU or where patients received lower levels of baseline respiratory support (3). A more recent non-randomised trial of 501 patients in the USA receiving supplemental oxygen for COVID-19 pneumonia and including non-ICU settings, reported worse outcomes in those allocated to APP, with patients requiring a higher level of oxygen support on day 5 (4). Differences in reporting of the APP intervention itself, patient compliance with the intervention and differences in comparator groups remain significant impediments to understanding these conflicting results. Nevertheless, better understanding is vital to inform decision-making, resource allocation and policy around APP, particularly in low-resource settings where limited staff already make implementing APP challenging (5,6).

In ventilated patients, duration of prone position is a key factor in determining its efficacy and data from APP studies also indicate that this is important (7). Accurately quantifying the duration of APP is difficult, particularly under pandemic pressures. Whilst several studies have not reported APP duration at all, others have relied on nursing reports, electronic health records or reports from patients themselves (5,8–14). In low-resource settings these methods are usually unfeasible or unavailable outside of ICUs. Duration may also be confounded by disease severity and implementation methods. More severely ill patients are more likely to be cared for in settings with greater access to staff for support in turning and maintaining APP, whereas patients with lower oxygen requirements, cared for in less intensive environments, are more able to move themselves to a position of their choice. Studies in the UK, Pakistan and China which have examined acceptability from patient perspectives suggest overall negative attitudes to APP related to discomfort, physical consequences and social factors which may influence patients’ willingness to self-prone (15–17). Concurrent patient perspectives from randomised controlled trials remains lacking.

Our aim in this study was to evaluate APP in an LMIC setting using a dedicated team to evaluate prone duration and assist participants with the allocated study intervention. Whilst conceived at the height of the pandemic in Vietnam, our study was implemented after widescale population-level vaccination coverage in Vietnam and is, to our knowledge, the only randomised trial in a largely vaccinated population. To remove the burden on staff we introduced wearable monitors to facilitate remote patient monitoring. Our team had already developed monitoring with low-cost pulse oximeters and in this trial wearables were used for both vital sign monitoring and quantification of prone position duration by incorporating a low-cost accelerometer (18).

## Methods

### Trial design

The study was an open-label randomized controlled trial. A Trial Steering Committee oversaw the trial and an independent Data and Safety Monitoring Review Board (DSMB) reviewed all severe adverse events and reviewed data for safety endpoints at pre-specified time-points. The full protocol is published separately. (19) The study was stopped before reaching the pre-determined sample size due to low case numbers and infeasibility of reaching the proposed sample size.

### Ethics

The protocol was approved by the Ethical Committee of the Hospital for Tropical Diseases and Oxford Tropical Research Ethics Committee. All participants or their representatives gave written informed consent before enrolment in the study.

### Setting

The study was carried out at the Hospital for Tropical Diseases, Ho Chi Minh City. The hospital is a tertiary referral centre for infectious diseases in southern Vietnam and was a designated special treatment centre for COVID-19 throughout the pandemic.

### Participants

Adult patients ≥18 years old with a diagnosis of probable or confirmed COVID-19 were eligible for inclusion to the study if they had moderate or severe COVID-19 (Vietnam Ministry of Health criteria, Supplementary materials) and required supplemental oxygen. Those already receiving non-invasive ventilation (continuous or bilevel positive airway pressure), mechanical ventilation, or with contraindication to prone positioning, body mass index > 35, pregnant, Glasgow coma scale <13 or a decision not to escalate care were excluded from the study. All participants or their representatives gave written consent prior to enrollment.

### Intervention

Those in the standard care group received verbal and written instructions conforming to the Vietnamese Ministry of Health Guideline as well as visits by the study team at the beginning and end of possible proning times. These instructions include changing position between prone, supine, lateral and semi-recumbent up every two hours (See Supplementary materials). Participants in the APP group were visited by the study team and given written and verbal advice about lying in the prone position as well as assistance with achieving and maintaining a fully prone position for as long as possible. All study procedures were carried out by a specific study team who were present in the ward 8am-5pm daily and dedicated ward nurses who supervised the evening APP session (6-8pm). Patients were followed daily for study outcomes until hospital discharge or transfer. Day 28 outcomes were collected by telephone follow-up for those participants already discharged from hospital at this time.

### Randomisation

Participants were enrolled by study staff prior to randomization. Enrolled patients were randomized in a 1:1 ratio to the two allocations according to computer-generated random list using block randomisation with variable block length to standard care or APP groups. An independent statistician generated this list.

### Procedures

APP was initiated as soon as possible after randomisation and continued until either escalation of therapy or cessation of oxygen therapy. Participants in the APP group were supported to be in the prone position for as long as possible while study staff were in attendance in the ward, excepting for mealtimes or other nursing procedures. Support included both physical assistance and physical aids such as pillows. No support was available to assist participants turning prone at night time after 8pm. Routine management was given by ward staff and followed Vietnam Ministry of Health Guidelines. In addition, all patients received continuous SpO_2_ (SmartCare Analytics, UK) and accelerometry (Axivity AX3, Newcastle upon Tyne, UK) monitoring with wearable devices during the intervention period. The pulse oximeters were connected via Bluetooth to a bedside tablet where data could be visualised, and also transmitted to cloud server for remote visualisation and downloading of data. (20). Accelerometers were attached to the patient’s infraclavicular fossa to infer prone vs non prone. Duration of prone position was also recorded manually by study staff in attendance in the ward. To determine acceptability of APP and wearable device monitoring, prior to hospital discharge, a questionnaire was administered to participants by trained study staff using a 10-point Likert scale to evaluate participant experiences (Supplementary materials (19)).

### Outcomes

The primary outcome was escalation of respiratory therapy within 28 days of randomisation, defined as intubation or escalation to next level respiratory support (with lowest level nasal canulae or face mask, escalating through HFNC to NIV or mechanical ventilation).

Secondary outcome measures included requirement for intubation and mechanical ventilation within 28 days of randomization, 28-day all-cause mortality, in-hospital mortality, duration of hospital stay, SpO_2_/respiratory rate/ heart rate/ FiO2 and ROX index ([SpO_2_/FiO_2_]/respiratory rate) – before and at end of period of prone positioning every day, duration of oxygen and estimated oxygen consumption. Daily monitoring for adverse events was performed by the study team.

A sample size was calculated based on treatment failure rate of 52%, a relative risk of treatment failure of 0.8 for the intervention, was calculated power at the two-sided 5% significance level, leading to 300 patients in each arm.

### Statistical Methods

Data analysis followed an a *priori* defined statistical analysis plan completed before database locking (See Supplementary Materials).

The primary analysis population for all analysis was the full population containing all randomized participants. Participants were analysed according to their randomized arm (intention-to-treat). Analyses for the primary endpoint were repeated on the per protocol population which excludes the following participants: participants not receiving the randomized intervention and other major violations of inclusion/exclusion criteria or study procedures.

The primary outcome measure, was compared between the groups based on a logistic regression model with the intervention as the only covariate. As odds ratios from logistic regression are somewhat difficult to interpret, we additionally estimated relative risk (RR) between the groups based on a binary regression model with a log-link rather than the logit link function used in logistic regression. The assessment of heterogeneity of the treatment effect was not performed because of small sample size. For secondary dichotomous outcomes such as requirement for intubation and mechanical ventilation within 28 days of randomisation, we computed the number of patients who developed or did not develop the outcomes of interest and fitted a logistic regression.

For duration of hospital stay, ventilator-free days, time from enrolment to first escalation of respiratory therapy, time from enrolment to first intubation, hospital mortality was treated as a competing event. Time-to-event analysis was performed using a subdistribution hazards model. Cause-specific cumulative incidence was estimated and plotted. Differences between intervention groups was tested using Gray’s log-rank test. In-hospital mortality was assessed with both a logistic regression model and estimated via Kaplan-Meier curves and compared using log-rank test. We assumed that individuals without death observed remained alive until day 90. People who were discharged home for palliative care were considered as in-hospital deaths.

For continuous outcomes such as vital signs (SpO_2_ /respiratory rate/ heart rate/ FiO_2_/ROX index) (repeated measurements) before and at end of prone sessions (morning and afternoon, excluding evening), interventions were compared using a linear mixed effects model, with or without a quadratic term for both fixed and random effect if it gives the model a better fit (p<0.05).

### Wearable data preprocessing

Accelerometers were worn continuously and oximeters during observation periods between 9am and 8am the following day. For accelerometry, 3 axis measures at 100 Hz frequency from the accelerometers were used to calculate the angular positions, defined by degrees in x-plane (left-right from -180 to 180), y-plane (prone-supine -180 to 180 degrees), and z-plane (up-down from -180 to 180). In order to determine ‘prone’ label for 30 second segments, we first smoothed the measures using median for 30 second consecutive moving window; then apply threshold of (|x| < 60, |y| < 40, |z| < 60) degrees. This threshold was obtained by grid search for |x|, |y| and |z| between 0 and 90, step 5 degrees, and optimised towards the prone morning and afternoon sessions observed by study staff.

### Adverse events

All adverse events, defined according to the Common Terminology Criteria for Adverse Events as “any untoward medical event that occurs to a study participant during the course of the study” and followed their grading (grade 1: mild to grade 4: severe) were recorded (19,21). Serious adverse events were defined as those which were life-threatening or resulted in death, new inpatient hospitalization or prolongation of existing hospitalization, persistent or significant disability or congenital anomaly. All serious adverse events and additional specified adverse events were reported to the study data monitoring and safety board and relevant ethical committees.

## Results

Ninety-three patients were enrolled between 8^th^ March 2022 and 23^rd^ March 2023 and followed up until 1^st^ May 2023 (Figure 1). Despite the Hospital for Tropical Diseases remaining the dedicated COVID-19 treatment centre for Ho Chi Minh City (population 10 million), a reduction in admissions with COVID-19 and eligible study participants meant recruitment was significantly impacted. The Trial Steering Committee and Data Monitoring and Safety board approved to early cessation of the study due to the unfeasibility of reaching the planned sample size, balanced with duty for timely reporting and sharing of already acquired data. Consequently 46 patients were enrolled in the APP group and 47 to standard care. Three participants were transferred to other wards before cessation of oxygen therapy and excluded from per-protocol analysis.

**Figure 1.**
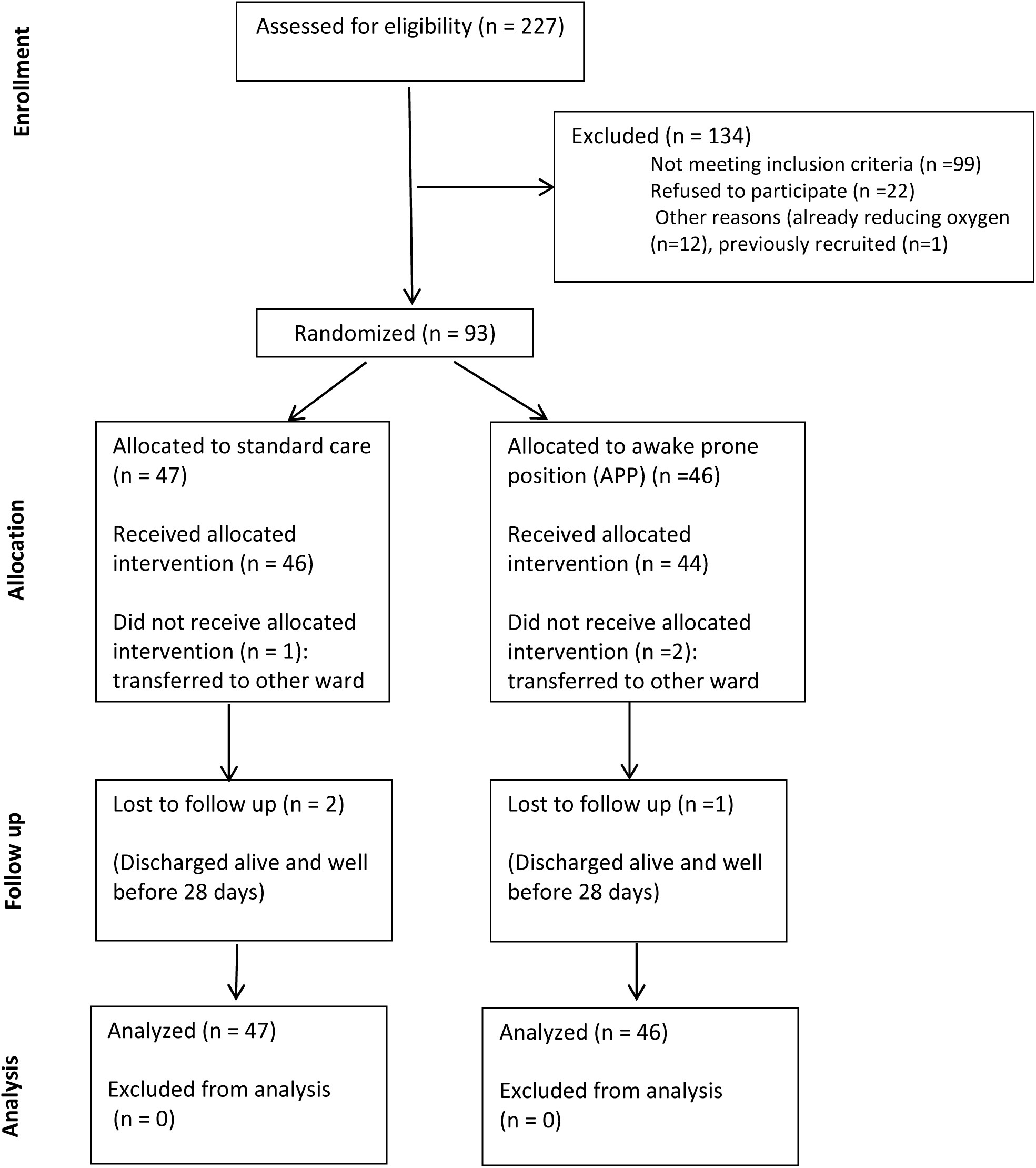
Study Flow Chart.

Baseline characteristics of participants are shown in Table 1. Eighty-five out of 93 (92%) of participants had received at least one dose of COVID-19 vaccine with 80/93 (86%) receiving 2 or more doses and 42/93 (45%) 3 or more doses respectively. Participants received the allocated intervention for a median 4.95 days (interquartile range (IQR) 3.0-7.8) in the standard care group and 3.94 days (IQR 2.9-7.2) in the APP group. Mean of daily APP duration per individual observed by study staff was a median of 0 hours (IQR 0.0, 1,2) in the standard care group and 3.3 hours (IQR 2.1, 4,7) in APP group.

**Table 1.**
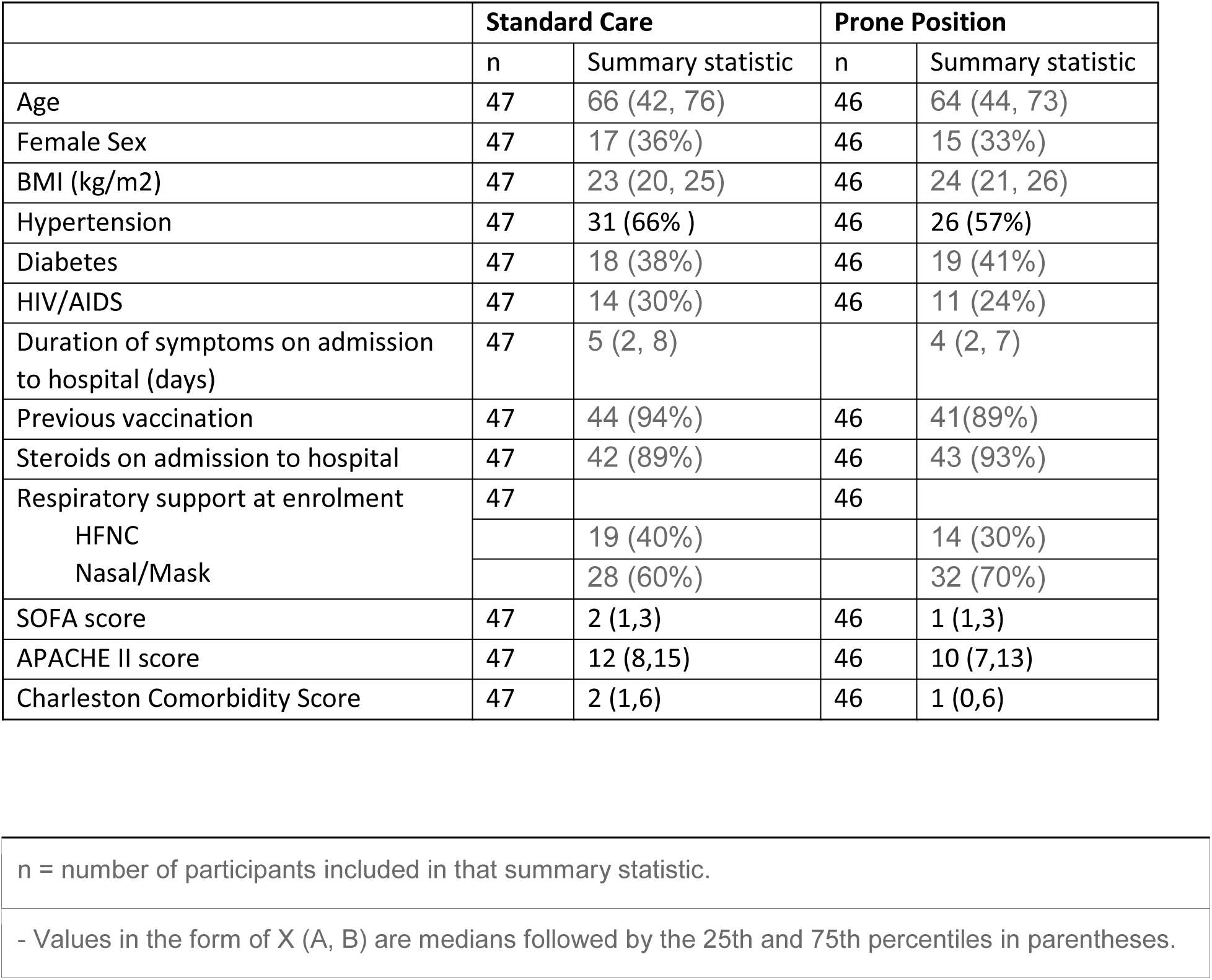
Baseline characteristics of participants.

There was no difference in the primary outcome of escalation of respiratory care within 28 days (RR 0.85, 95% CI 0.40, 1.78, p=0.67) (Table 2). For secondary outcomes of requirement for intubation and mechanical ventilation, 28-day mortality, in-hospital mortality, ventilator-free days and duration of oxygen therapy there was no difference between those allocated to APP or standard care (Table 3). Similarly we detected no changes in SpO_2_, respiratory rate, heart rate or ROX index before and after prone sessions (Table 3, Table S1 and Figure S1-S8).

**Table 2.** Primary outcome (intention to treat and per protocol population)

|  | Sample |  |  | Regression |  |  |
| --- | --- | --- | --- | --- | --- | --- |
|  | N | Non-escalation<sup>1</sup> | Escalatio<br>n<sup>1</sup> | RR<sup>2</sup> | 95%<br>CI<sup>2</sup> | p-value |
| **Intention to treat** | 93 |  |  |  |  |  |
| Standard care | 47 | 35 (74%) | 12 (26%) | — | — |  |
| Prone | 46 | 36 (78%) | 10 (22%) | 0.85 | 0.40, 1.78 | 0.67 |
| **Per protocol** | 90 |  |  |  |  |  |
| Standard care | 46 | 34 (74%) | 12 (26%) | — | — |  |
| Prone | 44 | 34 (77%) | 10 (23%) | 0.87 | 0.41, 1.82 | 0.71 |
| <sup>1</sup>n (%) |  |  |  |  |  |  |
| <sup>2</sup>RR = Relative Risk, CI = Confidence Interval |  |  |  |  |  |  |

**Table 3.** Secondary outcomes, intention to treat population.

|  | Standard care |  | APP |  | Outcome measure |  | P value |
| --- | --- | --- | --- | --- | --- | --- | --- |
|  | n | Summary statistic | n | Summary statistic |  | 95% CI |  |
| Intubation and MV within 28 days n (%) | 47 | 8 (17%) | 46 | 6 (13%) | RR 0.77 | 0.27, 2.04 | 0.59 |
| 28-day mortality n(%) | 47 | 8 (17%) | 46 | 8 (17%) | RR 1.02 | 0.41, 2.56 | 0.96 |
| In-hospital mortality n(%) | 47 | 11 (23%) | 46 | 10 (22%) | RR 0.93 | 0.43, 2.00 | 0.85 |
| Duration hospital stay (days) (median, IQR) | 47 | 12 (10, 20) | 46 | 11 (9, 16) | Beta -1.72 | -8.15, 4.72 | 0.60 |
| Duration of oxygen therapy<sup>1</sup> (days) median (IQR) | 47 | 6 (3,11) | 46 | 5 (3, 9) | Beta -1.32 | -3.48, 0.85 | 0.23 |
| Ventilation free days<sup>2</sup> (days) median (IQR) | 47 | 11 (9,16) | 46 | 10 (8, 15) | Beta -2.3 | -4.8, 0.2 | 0.07 |

1. Supplemental oxygen via low flow nasal canulae/mask or HFNC
2. Ventilation free days until hospital discharge

No participants experienced an adverse event attributed as definitely related to the intervention. There was no difference in adverse events judged as possibly related to the intervention groups (10/46 (24%) of standard care group and 8/44 (18%) of those allocated to APP. These events were pneumonia or exacerbation of chronic pulmonary disease (Table S2). Severe adverse events occurred in 9/44 (20%) participants in the prone group and 12/46 (26%) patients in the standard care group, p=0.56) (Table S3). None of these serious adverse events were judged as related to or possibly related to the intervention.

### Wearable device data

Data from wearable devices were available from all participants in whom devices had been used (45 in standard care and 44 in APP). Exploratory analyses showed increased monitoring data available from those in the standard care group, linked to longer duration of monitoring in those in the standard care group. Patient monitored in the standard care had gyrometry recording for median 118 hours (IQR 73, 187) compared to 93 hours (IQR 70, 171) in the APP group. Mean daily prone hours recorded were 4.3 hours (IQR 1.9, 7.4) in the standard care group compared to 7.3 hours (IQR 4.3, 9.2) in the APP group (p=0.006).

### Participant perspectives

Participant’s perspectives of APP revealed that most patients found APP comfortable. Similarly, they expressed general ease in getting into and out of the prone position (Table S4). Nevertheless, 29/43 (70%) participants in the APP group preferred to be in a supine position during the day and 23/43 (55%) preferred to be supine at night, which was similar to preferences reported by the standard care group. Participants reported the wearable monitoring devices very comfortable (median scores 8 (7 - 9) and 9 (8 - 9) out of 10 in standard care and prone groups respectively) (Table S4).

## Discussion

Our study terminated early due to a significant reduction in the number of patients hospitalised with COVID-19, reflecting the overall trend of the pandemic and success of the vaccination programme in Vietnam. Despite failing to reach the sample size, there are several important findings and conclusions.

Our final sample size was underpowered to detect the anticipated differences in outcomes and our study outcome rate was lower than expected perhaps due to vaccination status of our population and routine use of steroid treatment. On the other hand, we also did not detect significant differences in primary or secondary outcome measures. Nevertheless, we did not see any evidence of possible harm from the intervention. Importantly, in comparison to Qian et al(4) who reported high oxygen dependence at 5 days in patients treated with APP in a non-randomised clinical trial, we observed a trend towards shortening of supplemental oxygen therapy (nasal, mask or HFNC) in the APP group. This is further supported by the reduced monitoring hours in the APP group as monitors were removed when patients no longer required oxygen therapy.

The difference we observed in primary outcome between groups is similar to that reported in the large meta-analysis (3) and data from our study will be available to contribute to further analyses. Our study protocol and endpoints were deliberately designed to allow this. Of note is that our study is the first in a vaccinated population and the similarity in behavior of this population with previously studied unvaccinated patients underlines validity of our findings.

Importantly, our study has demonstrated that achieving significant duration of prone positioning in our population was extremely challenging, an important finding for policy-makers in LMICs such as Vietnam. Despite dedicated study staff available in the ward our average prone duration was less than our target 8 hours a day. Reasons for this included frequent interruptions to prone positioning due to routine ward care, mealtimes, as well as general frailty of our study population who required significant help to turn prone. We have demonstrated that APP is not a resource-free intervention. Staff are needed to communicate with patients, assist patients achieve and maintain the prone position and monitor for potential adverse events.

Our use of wearable devices and dedicated study staff allowed us to accurately quantify the time participants spent in the prone position. We note that accelerometer data indicates that participants in both standard and APP groups changed position frequently. The discrepancy between accelerometer recorded prone position and that observed by our study staff we believe is likely for several reasons.

Firstly, accelerometer data includes unobserved night-time movements of patients and secondly that our accelerometer data is the sum of 30 second intervals, thus may include many short periods of prone not accounted for by ward staff. Whilst our choice of the infraclavicular fossa for sensor positioning aimed to reduce false indication of position changing and focused our analysis on the position of the thorax, accelerometer data used arbitrary cut-offs and therefore thoracic positioning, not seen as ‘prone’ by observers may be classified as prone, for example side-lying.

We have demonstrated that applying technologies such as wearable devices may allow detailed monitoring to occur despite limited staff availability. Lack of routine electronic health record data and monitoring is a major impediment to carrying out high-quality clinical trials in LMICs without significant investment in trial staff and infrastructure. The use of novel technologies can potentially remove such barriers to participation redressing inequity in research and bias in data.

Our study improves the evidence base for APP as an intervention in COVID-19. Data will be included in an ongoing meta-analysis, emphasizing the value of harmonized outcome data and methodology used in ours and other studies. It remains unclear if the findings can translated to treatment of pneumonia due to other causes. Simple and effective means of improving outcomes of those with these infections could have important consequences for patients and resource-utilization.

## Data Availability

All data produced in the present study are available upon reasonable request to the authors

## Declarations

### Consent for publication

NA

### Availability of data and materials

the datasets used and/or analysed during the current study are available from the corresponding author on reasonable request.

### Authors’ contributions

Conceptualisation: CLT, LMY, GG, HBH; methodology NTP, NTN, LDVK, CTCV, NDT, DPT, NTT, VTL, TDK, AB, GET, DC,NTPD, EK, LVT; iImplementation: NTP, HBHm NTNm LDVK, LTTK, LHB, NTML, CTCV, DPT, NTDT, PTK, VTH, NTN, TTDV, JA, NTPD, EK, LVT, LMY; supervision: TTDV, NTP, NTN, TG, EK,NTPD, GET, LVT, NLNT, NTD; data collection & curation LTTK, LHBT, DPT, NTDT, PTK, VTH, NTN; analysis: DHD, HBH, LDVK, JA, CLT; manuscript drafting: DDH, HBH; review of manuscript: all authors.

## Acknowledgements

We would like to thank members of the DSMB, Nicholas White, Duncan Wyncoll, Bui Thi Hanh Duyen, Mavuto Mukaka; Trial Steering Committee: David Paterson, Mo Yin, Dang Trong Thuan, Vu Quoc Dat; patients and staff at the Hospital for Tropical Diseases, Ho Chi Minh City.

## References

1. Guérin C, Reignier J, Richard JC, Beuret P, Gacouin A, Boulain T, et al. Prone positioning in severe acute respiratory distress syndrome. N Engl J Med 2013 Jun 6;368(23):2159–68.

2. Ehrmann S, Li J, Ibarra-Estrada M, Perez Y, Pavlov I, McNicholas B, et al. Awake prone positioning for COVID-19 acute hypoxaemic respiratory failure: a randomised, controlled, multinational, open-label meta-trial. Lancet Respir Med. 2021;2600(21).

3. Li J, Luo J, Pavlov I, Perez Y, Tan W, Roca O, et al. Awake prone positioning for non-intubated patients with COVID-19-related acute hypoxaemic respiratory failure: a systematic review and meta-analysis. Lancet Respir Med. 2022;2600(22):7–9.

4. Qian ET, Gatto CL, Amusina O, Dear ML, Hiser W, Buie R, et al. Assessment of Awake Prone Positioning in Hospitalized Adults With COVID-19. JAMA Intern Med. 2022;

5. Fralick M, Colacci M, Munshi L, Venus K, Fidler L, Hussein H, et al. Prone positioning of patients with moderate hypoxaemia due to covid-19: Multicentre pragmatic randomised trial (COVID-PRONE). The BMJ. 2022;(February 2020).

6. Ibarra-Estrada M, Li J, Pavlov I, Perez Y, Roca O, Tavernier E, et al. Factors for success of awake prone positioning in patients with COVID-19-induced acute hypoxemic respiratory failure: analysis of a randomized controlled trial. Crit Care. 2022;26(1):1–13.

7. Barker J, Koeckerling D, Mudalige NL, Oyefeso O, Pan D. Awake prone positioning for patients with covid-19. The BMJ. 2022;(March):1–2.

8. Rosén J, von Oelreich E, Fors D, Jonsson Fagerlund M, Taxbro K, Skorup P, et al. Awake prone positioning in patients with hypoxemic respiratory failure due to COVID-19: the PROFLO multicenter randomized clinical trial. Crit Care. 2021;25(1):1–10.

9. Ehrmann S, Li J, Ibarra-Estrada M, Perez Y, Pavlov I, McNicholas B, et al. Awake prone positioning for COVID-19 acute hypoxaemic respiratory failure: a randomised, controlled, multinational, open-label meta-trial. Lancet Resp Med. 2021;9:1387–95.

10. Alhazzani W, Parhar KKS, Weatherald J, Al Duhailib Z, Alshahrani M, Al-Fares A, et al. Effect of Awake Prone Positioning on Endotracheal Intubation in Patients with COVID-19 and Acute Respiratory Failure: A Randomized Clinical Trial. In: JAMA - Journal of the American Medical Association. American Medical Association; 2022. p. 2104–13.

11. Jayakumar D, Ramachandran, DNB P, Rabindrarajan, DNB E, Vijayaraghavan, MD BKT, Ramakrishnan, AB N, Venkataraman, AB R. Standard Care Versus Awake Prone Position in Adult Nonintubated Patients With Acute Hypoxemic Respiratory Failure Secondary to COVID-19 Infection—A Multicenter Feasibility Randomized Controlled Trial. J Intensive Care Med. 2021;36(8):918–24.

12. Rampon G, Jia S, Agrawal R, Arnold N, Martín-Quirόs A, Fischer EA, et al. Smartphone-Guided Self-prone Positioning vs Usual Care in Nonintubated Hospital Ward Patients With COVID-19: A Pragmatic Randomized Clinical Trial. Chest. 2022 Oct 1;162(4):782–91.

13. Kharat A, Dupuis-Lozeron E, Cantero C, Marti C, Grosgurin O, Lolachi S, et al. Self-proning in COVID-19 patients on low-flow oxygen therapy: a cluster randomised controlled trial. ERJ Open Res. 2021;7(1):00692–2020.

14. Johnson SA, Horton DJ, Fuller MJ, Yee J, Aliyev N, Boltax JP, et al. Patient-directed prone positioning in awake patients with COVID-19 requiring hospitalization (PAPR). Ann Am Thorac Soc. 2021;18(8):1423–6.

15. Sethi SM, Hirani S, Iqbal R, Ahmed AS. Patient’s Perspective of Awake Proning: A Cross-Sectional Interview-Based Survey from COVID-19-Recovered Patients. Crit Care Explor. 2022 Dec 16;4(12):E0824.

16. Jha A, Chen F, Mann S, Shah R, Abu-Youssef R, Pavey H, et al. Physiological effects and subjective tolerability of prone positioning in COVID-19 and healthy hypoxic challenge. ERJ Open Res. 2022 Jan 1;8(1).

17. Zhu L, Ni Z, Zhang Y, Zhan Y, Lan M, Zhao R. Barriers and facilitators of adherence to awake prone positioning: a qualitative study using the COM-B model. BMC Pulm Med. 2023 Dec 1;23(1).

18. Chau NVV, Hai HB, Greeff H, Phan Nguyen Quoc K, Trieu HT, Khoa LDi Van, et al. Wearable remote monitoring for patients with COVID-19 in low-resource settings: Case study. BMJ Innov. 2021;7:S12–5.

19. Truong NT, Phong NT, Nguyen NT, Khanh LTT, Tran LHB, Linh NTM, et al. Evaluation of awake prone positioning effectiveness in moderate to severe COVID-19. Wellcome Open Res. 2023 Jun 5;8:235.

20. Chau NVV, Trung TN, Nguyen P, Khanh Q, Tran P, Nhat H, et al. Wearable devices for remote monitoring of hospitalized patients with COVID-19 in Vietnam. Wellcome Open Res. 2022;1–7.

21. Common Terminology Criteria for Adverse Events [Internet]. Available from: http://ctep.cancer.gov/protocolDevelopment/electronic_applications/ctc.htm

