## Supplementary material for "Awake prone positioning effectiveness in moderate to severe COVID-19 a randomized controlled trial"

### Supplementary Materials:

#### Study outcomes

##### Table S1: Secondary outcomes, per protocol population

| | Standard care | | Prone | | Outcome measure | | P value |
|---|---|---|---|---|---|---|---|
| | n | Summary Statistic | n | Summary Statistic | | 95% CI | P value |
| Intubation and MV within 28 days n(%) | 46 | 8 (17%) | 44 | 6 (14%) | RR 0.78 | 0.28, 2.08 | 0.62 |
| 28-day mortality n(%) | 46 | 8 (17%) | 44 | 8 (18%) | RR 1.05 | 0.42, 2.61 | 0.92 |
| In-hospital mortality n(%) | 46 | 11 (24%) | 44 | 10 (23%) | RR 0.95 | 0.44, 2.04 | 0.89 |
| Duration hospital stay (days) (median, IQR) | 46 | 13 (10, 21) | 44 | 11 (9, 16) | Beta -1.73 | -8.36, 4.91 | 0.61 |
| Duration of oxygen therapy[1] (days) (median, IQR) | 46 | 6 (3,7) | 44 | 5 (3,6) | Beta -1.25 | -3.48, 0.97 | 0.27 |
| Ventilator free days[2] (median, IQR) | 46 | 11 (9,17) | 44 | 9 (8,15) | Beta -2.41 | -5.0, 0.2 | 0.07 |

1. Supplemental oxygen via low flow nasal canulae/mask or HFNC

2. Ventilation free days until hospital discharge

#### Figure S1. Time to death according to intervention group – intention to treat population

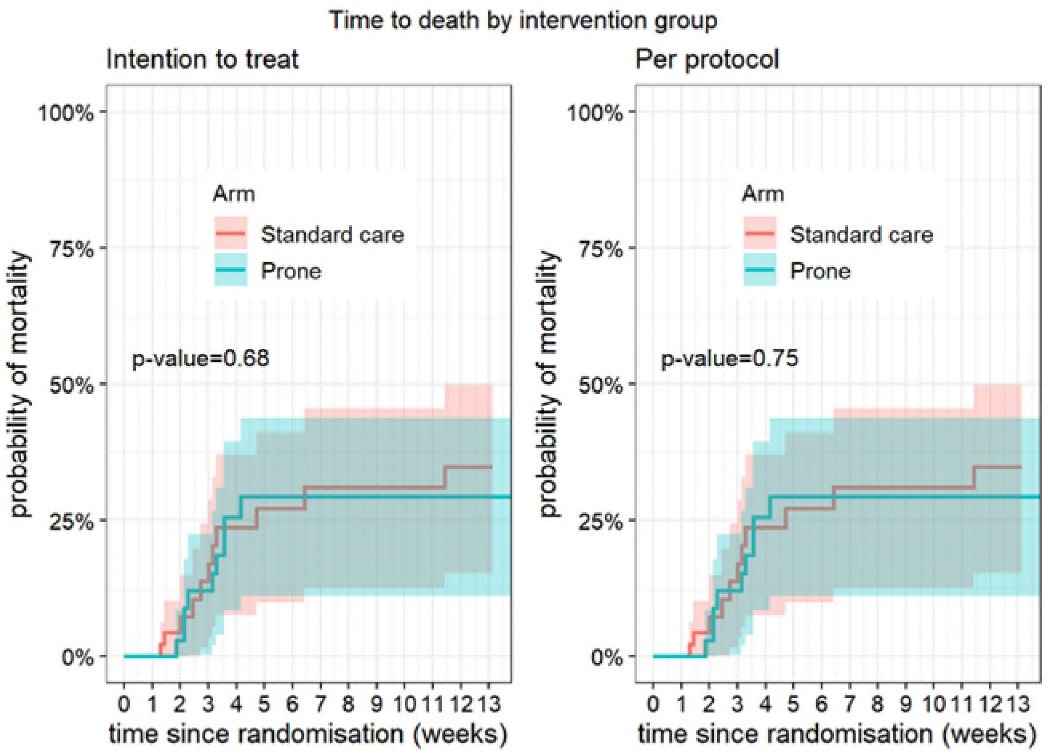

#### Figure S2 Time to hospital discharge (intention-to-treat and per-protocol populations)

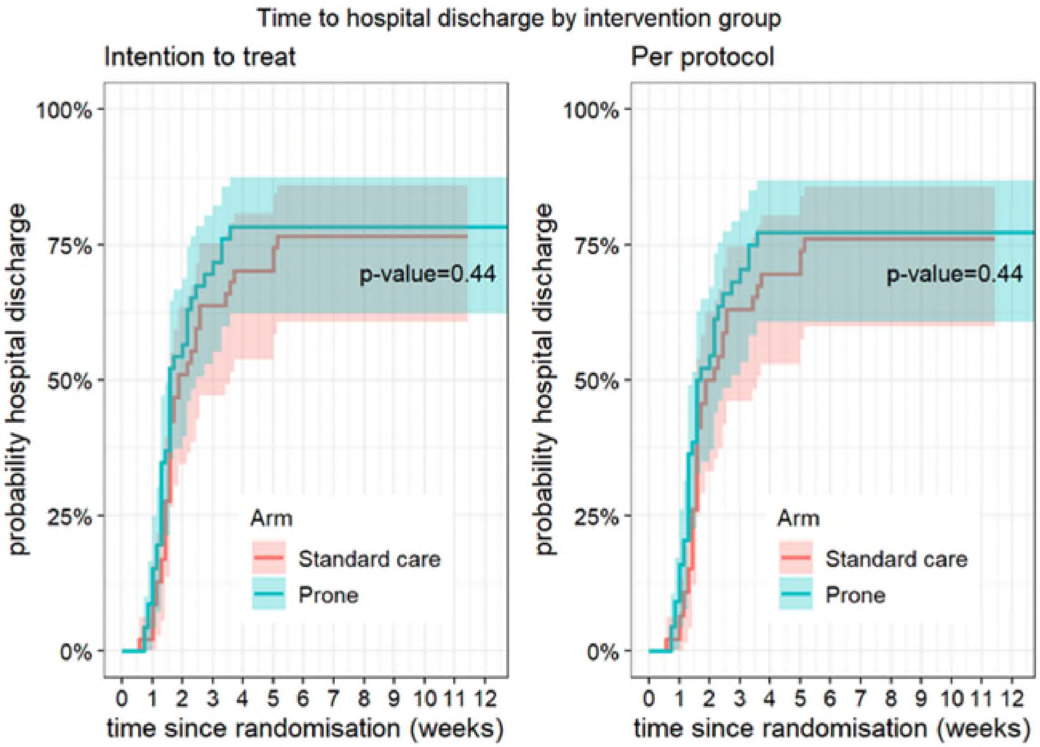

P value relates to cause-specific cumulative incidence tested using Gray’s log-rank test

#### Figures S3 Kaplan Meier Curves for mortality 28-day all-cause mortality (intention-to-treat and per-protocol populations)

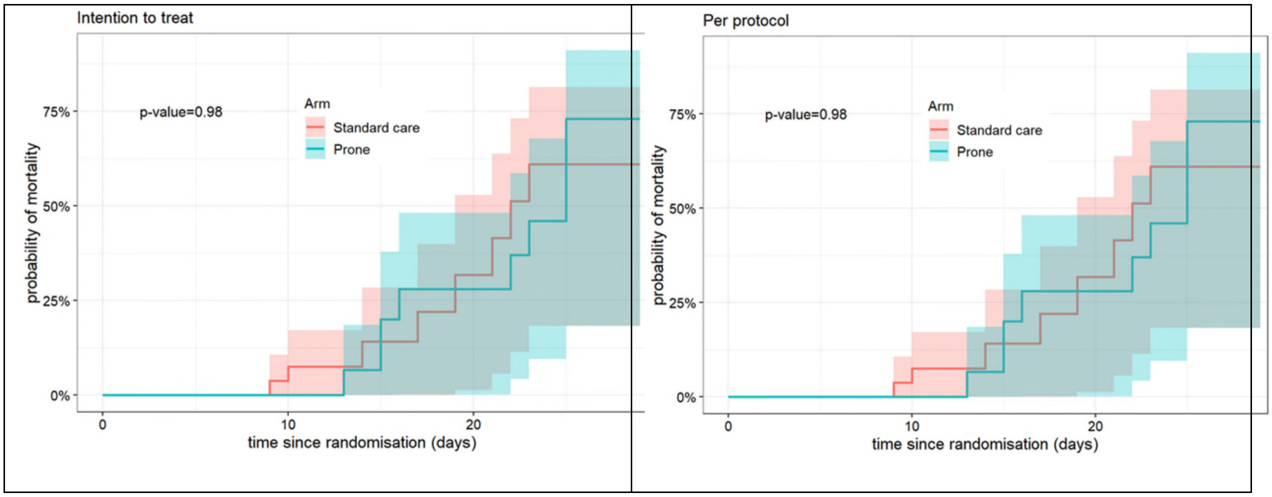

P value relates to cause-specific cumulative incidence tested using Gray's log-rank test

#### Figure S4 Longitudinal vital signs parameters: $SpO_2$ (intention-to-treat and per-protocol populations)

Intention to treat

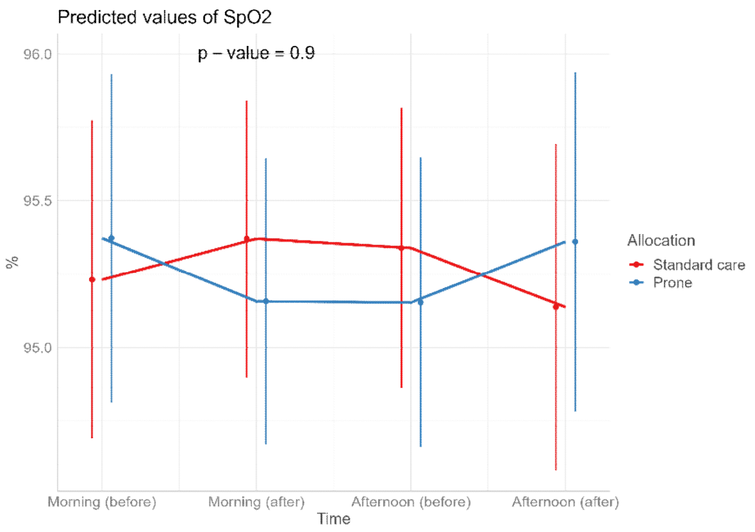

Per protocol

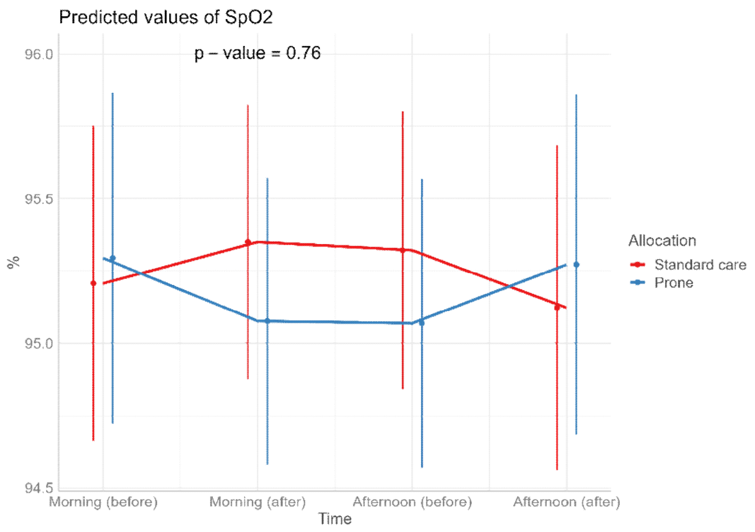

The p-values refer to the overall effect of intervention on the outcome

#### Figure S5 Longitudinal vital signs parameters: respiratory rate (intention-to-treat and per-protocol populations)

Intention to treat

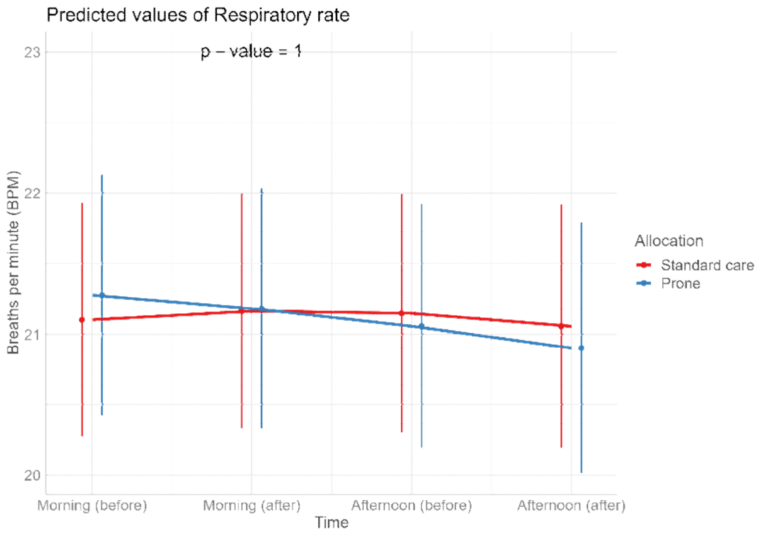

Per protocol

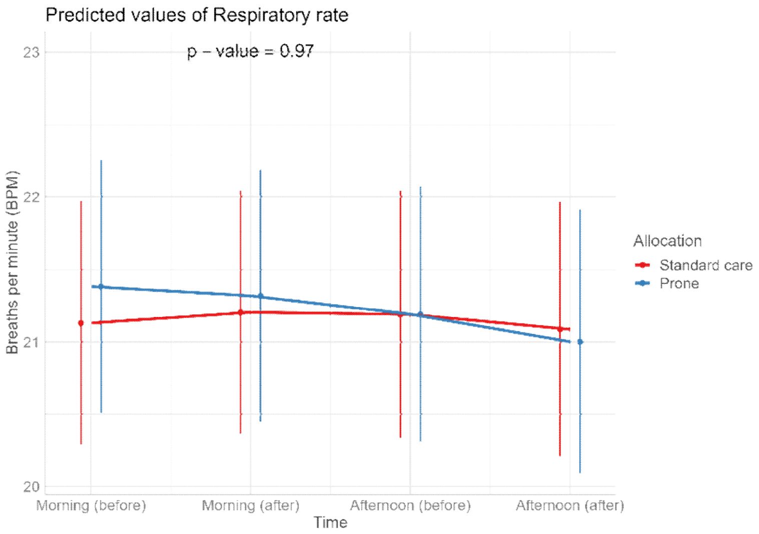

The p-values refer to the overall effect of intervention on the outcome

#### Figure S6 Longitudinal vital signs parameters: heart rate (intention-to-treat and per-protocol populations)

Intention to treat

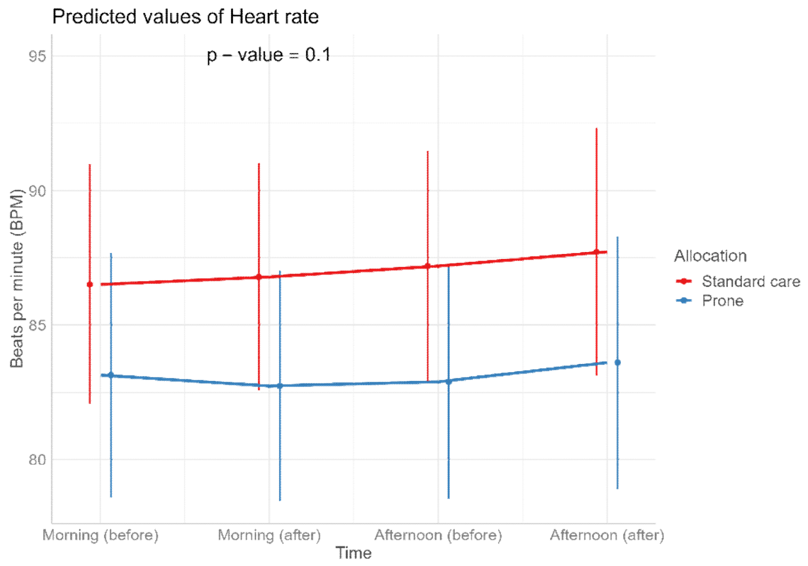

Per protocol

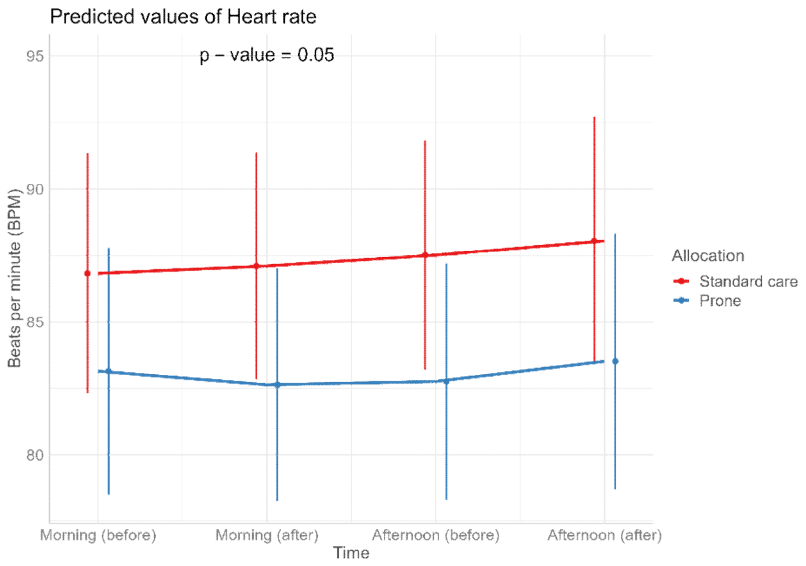

The p-values refer to the overall effect of intervention on the outcome

#### Figure S7 Longitudinal vital signs parameters: $FiO_2$ (intention-to-treat and per-protocol populations)

Intention to treat

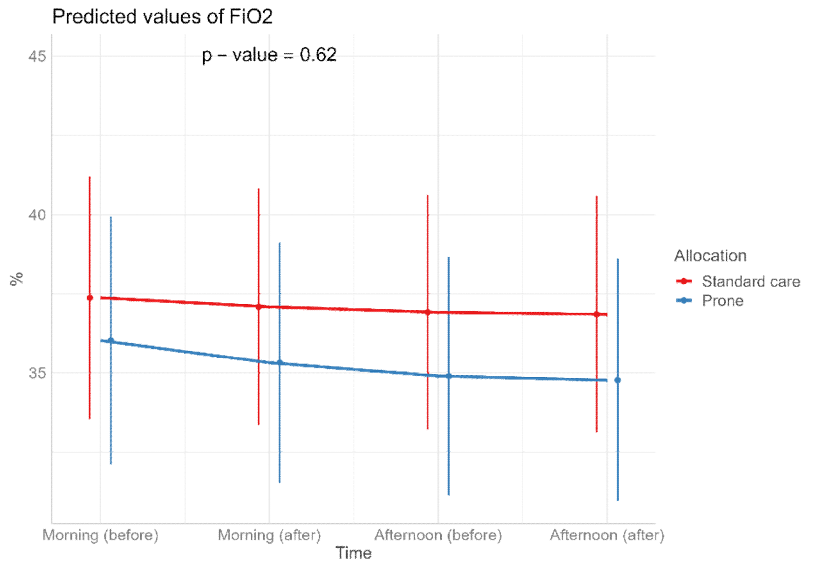

Per protocol

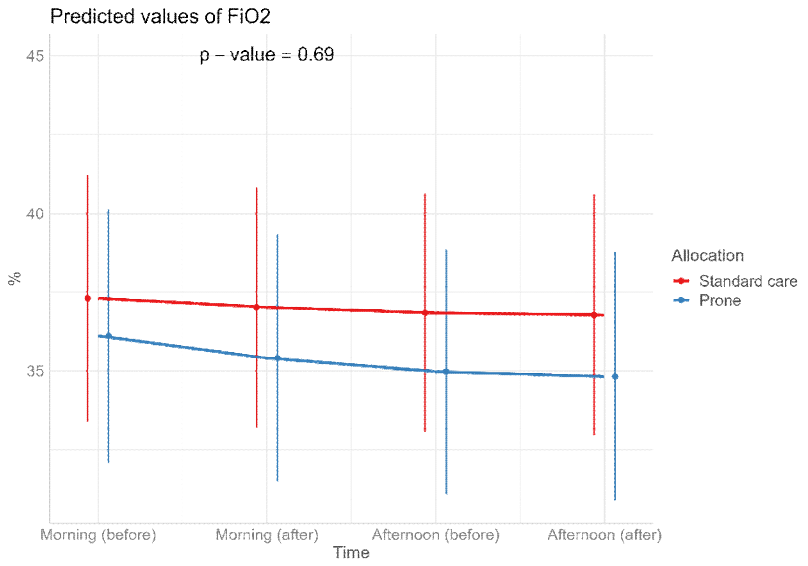

The p-values refer to the overall effect of intervention on the outcome

#### Figure S8 Longitudinal vital signs parameters: ROX (intention-to-treat and per-protocol populations)

Intention to treat

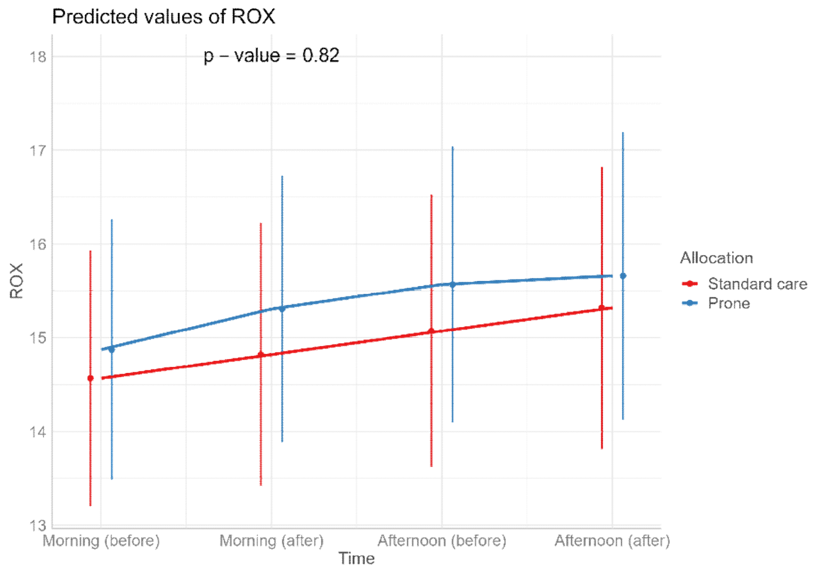

Per protocol

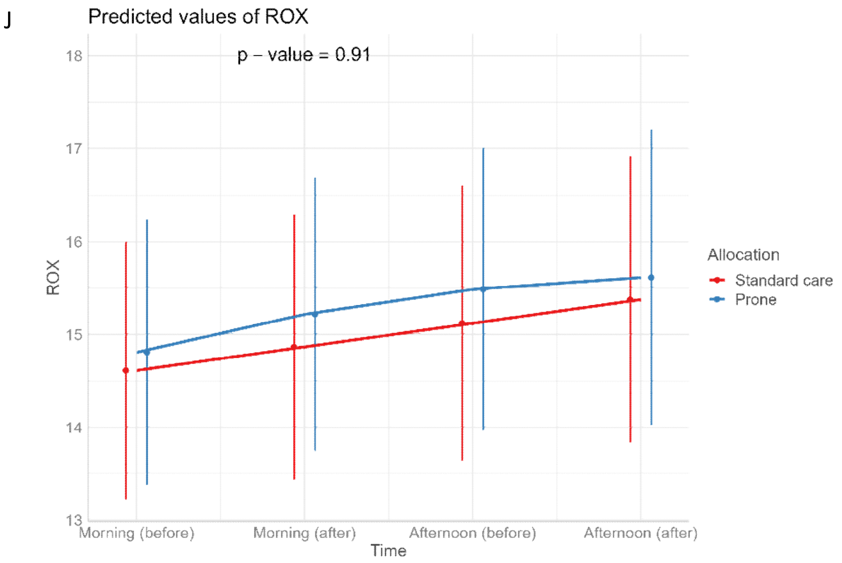

The p-values refer to the overall effect of intervention on the outcome

#### Table S2. Adverse events, per protocol population

| | Prone (N=44) | | STDCARE (N=46) | | |
|---|---|---|---|---|---|
| **Characteristic** | **n** | **Summary statistic** | **n** | **Summary statistic** | |
| Definitely Related AE | 44 | 0/44 (0%) | 46 | 0/46 (0%) | - |
| Possibly Related AE | 44 | 8/44 (18%)* | 46 | 10/46 (22%)** | 0.673 |
| Number of AEs per patient | 44 | | 46 | | 0.624 |
| - 0 | | 7/44 (16%) | | 3/46 (7%) | |
| - 1 | | 4/44 (9%) | | 5/46 (11%) | |
| - 2 | | 3/44 (7%) | | 7/46 (15%) | |
| - 3 | | 6/44 (14%) | | 6/46 (13%) | |
| - 4 | | 6/44 (14%) | | 5/46 (11%) | |
| - 5 | | 5/44 (11%) | | 3/46 (7%) | |
| - >5 | | 13/44 (30%) | | 17/46 (37%) | |

*7 pneumonia, 1 blood stream infection/pneumonia; **10 pneumonia or exacerbation of chronic pulmonary disease

#### Table S3. Severe adverse events, per protocol population

| | Prone (N=44) | | Standard care (N=46) | | |
|---|---|---|---|---|---|
| **Characteristic** | **n** | **Summary statistic** | **n** | **Summary statistic** | |
| Severe Adverse Event | 9 | | 12 | | 0.56 |
| - Cardiopulmonary Failure | | 1/9 (0%) | | 1/12 (8%) | |
| - Multi organ failure, shock | | 6/9 (67%) | | 6/12 (50%) | |
| - Multiorgan failure | | 0/9 (0%) | | 2/12 (17%) | |
| - Septic shock | | 1/9 (11%) | | 3/12 (25%) | |

#### Table S4. Questionnaire Results

| | Standard Care (N=42) | | Prone (N=43) | |
|---|---|---|---|---|
| Comfort of prone position (0-10 scale, with 0 extremely uncomfortable and 10 extremely comfortable), median (1st and 3rd quartile) | n=21 | 8 (6, 8) | n=42 | 7 (6, 8) |
| Ease of getting in and out of prone position (0-10 scale, with 0 very difficult and 10 very easy), median (1st and 3rd quartile) | n=21 | 8 (5.5, 9) | n=42 | 8 (6, 8) |
| Comfort of wearable devices ((0-10 scale, with 0 extremely uncomfortable and 10 extremely comfortable), median (1st and 3rd quartile) | n=43 | 8 (7, 9) | n= 42 | 9 (8, 9.25) |
| Preferred daytime position whilst receiving<br>oxygen n (%)<br>Supine<br>Prone<br>Side<br>Sitting up<br>No preference | n=43 | <br>30 (70)<br>3 (7)<br>6 (14)<br>4 (9)<br>0 | n=42 | <br>29 (70)<br>3 (7)<br>8 (19)<br>2 (5)<br>0 |
| Preferred nighttime position whilst receiving<br>oxygen n( %)<br>Supine<br>Prone<br>Side<br>Sitting up<br>No preference | n=43 | <br>24 (56)<br>1 (2)<br>18 (42)<br>0 (0)<br>0 (0) | n=42 | <br>23 (55)<br>2 (5)<br>16 (38)<br>0 (0)<br>1 (2) |

#### Statistical Analysis Plan for "Awake prone positioning in moderate to severe COVID-19: a cluster randomized controlled trial".

| Version Number & date: 1.0 10th March 2023 | | | |
|---|---|---|---|
| **Author** | **Position** | **Signature** | **Date** |
| Duc Du | Trial Statistician | | |
| Louise Thwaites | OUCRU PI | | 5/24/23 |

**Revision History**

| Version | Author | Date | Reason for Revision |
|---|---|---|---|
| 1.1 | Duc Du | 10.3.23 | Additional background description on title and introduction pages |
| 1.2 | Louise Thwaites | 28.5.23 | Adjustment variable names given for some outcome variables |
| 1.3 | Louise Thwaites | 22.6.23 | Variable field names added for CCI and vaccination status (baseline characteristics) |
| 1.4 | Louise Thwaites | 27.9.23 | Variable field names for AE updated. Note that separate file for AE"s is included in current datasheet. |
| 1.5 | Louise Thwaites | 15.10.23 | Corrected GCS typo and A-a calculation APACHE score |
| 1.6 | Louise Thwaites | 11.01.24 | Clarified comorbidity and APACHE score |
| 1.7 | Louise Thwaites | 27.3.24 | Typo in Comorbidity column and SOFA |

This analysis plan was written by Duc Du and Louise Thwaites

This document details the final analysis for the cluster randomised controlled clinical trial OUCRU 06NV conducted at the Hospital for Tropical Diseases, Ho Chi Minh City as outlined in the trial protocol version 1.1 September 2022. The study was a superiority trial investigating whether prone positioning of hospitalized Vietnamese patients with moderate to severe COVID-19 for ≥ 8 hours a day reduces treatment failure. This document contains detailed definitions of the endpoints. Deviations from the original protocol are noted.

#### DATA SOURCES

The data-source for this analysis is the CliRes study database 06NV which contains multiple tables (e.g. **ENR** contains patients' enrolment information, **OUTCOME SUMMARY** contains treatment outcome (e.g. hospital mortality, escalation to next level respiratory support history etc). The study Data Management Plan, Standard Operating Procedures include further information.

In this analysis plan, we refer to variables within tables by separating them by a dot, e.g. **ENR**. ALLOCATION refers to the variable ALLOCATION in table **ENR** and indicates the treatment arm "Standard care" or "Prone" that a patient was randomised.

#### TRIAL DESIGN AND SAMPLE SIZE

##### Trial design

Pragmatic open label randomised controlled trial.

##### Sample size

Our sample size is based on local data showing a current treatment failure rate of approximately 52%, a relative risk of treatment failure estimated at 0.8 for the intervention, corresponding to an absolute risk reduction from 52% to 40 %. To detect this reduction with 80% power at the two-sided 5% significance level, 287 patients are required in each arm, giving a total sample size of 580 allowing for exclusions and withdrawals.

Since the trial was unexpectedly stop before completed due to low rate of hospitalisation, the final sample size was deviated. Giving total sample size of 93 recruited for both arms (46 each arm), we can detect the proposed expected reduction rate of treatment failure at 0.8 for the intervention with 20% power at the two-sided 5% significance level.

The given final total sample size of 93 recruited for both arms (46 each arm) also allows to detect a reduction rate of 0.5 for the intervention with 80% power at the two-sided 5% significance level.

| Study | Power | p1 | p2 | Effect size (Cohen's d) | n1 | n2 | N |
|---|---|---|---|---|---|---|---|
| 1 | 0.8 | 0.52 | 0.40 | 0.24 | 273 | 273 | 546 |
| 2 | 0.8 | 0.52 | 0.42 | 0.21 | 356 | 356 | 712 |
| 3 | 0.8 | 0.46 | 0.40 | 0.12 | 1090 | 1090 | 2180 |
| 4 | 0.8 | 0.82 | 0.32 | 1.06 | 14 | 14 | 28 |
| Final sample size | | | | | | | |
| | 0.2 | 0.52 | 0.40 | 0.24 | 43 | 43 | 86 |
| | 0.8 | 0.52 | 0.26 | 0.54 | 42 | 42 | 84 |

#### ANALYSIS POPULATIONS

There are two main populations defined:

1. **The intention to treat (ITT) population** consists of all patients who have been randomised to the trial (derived as **ENR**.ENRDTC with a date, i.e. not missing data). Analysis will be according to the randomized treatment arm (from randomization list).
2. **The per-protocol population (PP)** consists of all patients who received the allocated intervention. Excluded individuals will be supplied as a separate list. Analysis will be according to the randomized treatment arm (from randomization list).

#### STUDY FLOW AND COMPLETENESS OF FOLLOW-UP

We will summarize the following quantities related to follow-up duration by treatment arm:
Number screened (Supplied on separate list); Reasons for ineligibility (supplied as separate list)

Number randomised (**ENR**.ENRDTC not blank)
Number of subjects who completed of study in hospital
Number of subjects who completed 28 days follow up after randomization

Number of subjects who withdrew from study

#### PRIMARY ENDPOINTS

Escalation of respiratory therapy or intubation within 28 days of randomization, defined as any of the following:

- Escalation to next level respiratory support (with lowest level nasal canulae or face mask
  - (**ESC.**Level1 = "nasal" or "mask", escalating through HFNC to NIV or mechanical ventilation).
  - OR
  - **ESC**.ESCULATE = "YES"
- Intubation
  - **ESC.**Level1, ESC. Level 2, ESC.Level3, ESC.Level4 or ESC.Level5 ="MV")

#### SECONDARY ENDPOINTS

- Requirement for intubation and mechanical ventilation within 28 days of randomization
  - **(ESC.** ESCULATE = "YES" an**d** , ESC. Level 2, ESC.Level3, ESC.Level4 or ESC.Level5 ="MV" AND **ENR.**ADMDTC **ESC.**1st_ESC_DATE <28)
- 28-day all-cause mortality
  - (**ESC.** D28_STATUS = "Death").
- Hospital mortality
  - **ESC**. DATE_DEATH !=""
- Duration of hospital stay
  - (**ENR.**ADMDTC

**R_DISC.** DATE_OF_HOSPITAL_DISCHARGE).

- Duration of intensive care unit stay
  - ( **R_DISC.**DATE_OF_DISCHARGE - **ENR.**ADMDTC )
- $SpO_2$ /respiratory rate/ heart rate/ $FiO_2$ and FiO2:SpO2 – before and at end of period of prone positioning every day . exclude evening values
  - (**DAILY.**M_SPO2ARV - **DAILY.**M_SPO2END) AND (**DAILY**.A_SPO2ARV - **DAILY**.A_SPO2END)
  - **(DAILY**.M_RESPARV - **DAILY**.M_RESPEND) AND **(DAILY**.A_RESPARV - **DAILY**.A_RESPEND)
  - **(DAILY**. M_HRARV - **DAILY**. M_HREND) AND **(DAILY**. A_HRARV - **DAILY**. A_HREND)
  - **(DAILY**.M_O2ARV, **DAILY**.M_O2END) AND **(DAILY**. A_O2ARV **DAILY**.M_O2END)
  - (**DAILY.**M_SPO2ARV/**DAILY**.M_O2ARV) -

(**DAILY.**M_SPO2END/**DAILY**.M_O2END) AND (**DAILY**.A_SPO2ARV/**DAILY**.A_O2ARV) - (**DAILY**.A_SPO2END)/ **DAILY**.A_O2END)

- ROX index (ratio of $SpO_2$: $FiO_2$ to respiratory rate) before and at end of period of prone positioning every day
  - (**DAILY.**M_SPO2ARV/**DAILY.**M_O2ARV/**DAILY.**M_RESPARV) -

(**DAILY.**M_SPO2END/**DAILY.**M_O2END/**DAILY.**M_RESPEND) AND

(**DAILY**.A_SPO2ARV/**DAILY**.A_O2ARV/**DAILY**.A_RESPARV) - (**DAILY**.A_SPO2END)/ **DAILY**.A_O2END) **DAILY**.A_RESPEND)

- Duration of oxygen therapy
  - (**R.SRDAILY**.FRACTION_INSPIRED_OXYGEN > 0.21 AND **R.SRDAILY**.MECHANICALLY_VENTILATED = "self_vent")
- Ventilator free days
  - Duration of hospital stay - (**R.SRDAILY.**MECHANICALLY_VENTILATED **=≠ "**mechanical_vent"
- Time from enrolment to first escalation of respiratory therapy
  - [**ESC**.L1_Date_time - **ENR.**ENRDTC for those with escalation]
- Time from enrolment to first intubation
  - **ESC.**1st_ESC_DATE - **ENR.**ADMDTC]
- Adverse events –Supplied separately – adverse events possibly or probably related to intervention.

**AE_AE**.AE_YES_No = "YES" described by **AE_AE**.Type. AE_AE.GRADE, **AE_AE**.REALTIONSHIP

- SAEs described by **AE_SAE**.Actual SAE = 1 AND described by **AE_SAE.**Event and **AE_SAE.** SAE_Relationship
- Oxygen consumption (estimated from flow rate and ventilation method) - money (**ESC.** O2_FEE_TOTAL)
- Duration of prone position [median hours/day during eligible proning time
  - **DAILY**.EPIN24HR for days where:

**DAILY**.M_TIMEARV, **DAILY**.A_TIMEARV, **DAILY**. E_TIMEARV **≠ " "** AND

**(DAILY.**M_OXYGENARV **DAILY.**A_OXYGENARV **DAILY.** E_OXYGENARV ≠ "AIR " or "MV")

- Additional endpoints, measured in this group of patients but reported separately, include acceptability from patients' perspectives and hospital direct medical costs.

### ANALYSIS

#### Primary endpoint

The main analysis is the comparison of escalation respiratory therapy or intubation within 28 days of randomization (binary variable: YES/NO) as defined between prone position intervention versus standard care. Patients will be analysed according to their randomized arm. All inference is based on likelihood ratio tests. All effect measures will be supplied with p-values and 95% confidence intervals.

Escalation of respiratory therapy or intubation within 28 days of randomization is summarized as x/n (%) in each treatment group and compared between the groups based on a logistic regression model with the intervention as the only covariate (prone position intervention versus standard care). As odds ratios from logistic regression are somewhat difficult to interpret, we will additionally estimate relative risk between the groups based on a binary regression model with a log-link rather than the logit link function used in logistic regression.

We will assess heterogeneity of the treatment effect by including interactions with the following variables (one at a time, not all together in a single model) and report the odds ratios for both treatment arms and covariates, irrespective of statistical significance of the interaction.

- Age (**ENR**.AGE), using restricted cubic splines with three knots.
- Other covariates.

For the numeric variables, we plot the odds ratios for the range of the numeric variable in a figure for both treatment arms.

#### Secondary endpoints

For requirement of intubation and mechanical ventilation within 28 days of randomisation as defined (binary variable: YES/NO), analysis will be done the same with primary endpoint to compare between prone position intervention versus standard care.

For duration of hospital stay, duration of intensive care unit (ICU) stay, duration of ventilation, time from enrolment to first escalation of respiratory therapy, time from enrolment to first intubation and duration of supplemental oxygen free days, hospital mortality is treated as a competing event. Those that were discharged palliatively are considered as in hospital deaths as well and are assumed to have died at discharge. For duration of ventilation, time from enrolment to first escalation of respiratory therapy and time from enrolment to first intubation, people who have ventilation or respiratory therapy or intubation stopped to allow them to go home to die will be considered as deaths as well. So, if end of ventilation day/ end of respiratory therapy / end of intubation = day of death or palliative discharge then we should consider this as a death for analysis. We will nonparametrically estimate the cause-specific cumulative incidence for each of the three event types and death, and we plot the results. We test for differences between the treatment groups using Gray's log-rank test. Contrary to the protocol, no Cox regression is performed.

For 28-day all-cause mortality, hospital mortality, we compute the number of patients as x/n (%) for each outcome in each treatment group and compared between the groups based on a logistic regression model. Mortality up to 28 days and hospital mortality will also be visualised using Kaplan-Meier curves and treatment intervention will be compared using the log-rank test.

Daily $SpO_2$ /respiratory rate/ heart rate/ $FiO_2$/FiO2:SpO2 or ROX index (repeated measurements every 5 minutes) before and at end of periods (morning and afternoon, excluding evening) of prone positioning every day will be compared using a linear mixed effects model. We will consider a linear trend for fixed and random effect, but will add a quadratic term for both effects if it gives a better model fit ($p<0.10$).

The cumulative amounts of oxygen consumption and duration of prone position may have a skewed distribution. Therefore, we use the Box-Cox procedure with intervention arm as covariable to find a suitable transformation. If reasonable, we use the identity or log transformation. After the transformation, arms are compared using linear regression. As effect measure we report the difference in expected value on the transformed scale (and report the transformation we used). Since this may be hard to interpret, we also plot the distribution of the cost variable by intervention arm via histograms. We add to the histograms the mean value with 95% confidence interval for each intervention arm.

#### Adverse events

We consider "any adverse event" as well as each adverse event (AE) separately. Tables will be generated to summarize the proportion of individuals with the adverse event, tabulating adverse events by grade (I-IV) and whether these events were judged to be related or possibly related to the treatment intervention [**AE_AE.**AE_YES_NO ="YES" **AE_AE.**Type**.**, **AE_AE.**Grade**AE_AE**.RELATIONSHIP , "POSREL" or "REL"].

Tables will be generated separately for severe adverse events [***AE.SAE*** *EVENT* ]. Comparisons of the proportions will be done with the chi-square test for independence; if the expected number is <= 1 in at least one of the cells, Fisher's exact test is used.AE and SAE data will be supplied part of exported dataset 28.5.23

#### OTHER DESCRIPTIVE ANALYSES

##### Summary of baseline characteristics

Baseline characteristics will be summarized as median (1st and 3rd quartile, lowest and highest value) for numeric data and n (%) for categorical data. No formal statistical comparison of baseline characteristics between the two study arms will be performed.

The following baseline characteristics will be summarized:

a. Patient details:

- Sex, age (**ENR.**AGE, **ENR.**SEX)),
- body mass index (**R_SRASMT**.WEIGHT/ (**R_SRASMT**.HEIGHT)$^2$ ),
- Charlson Comorbidity Index (see appendix),
- Hypertension (**R.ADM.**CORMBID_CONDITIONS1/2/3/4/OTHER = "Hypertension",
- Diabetes (**R.ADM.**CORMBID_CONDITIONS1/2/3/4/OTHER = "Type 1 diabetes", "Type 2 diabetes", "Type 1 diabetes with complications" or "Type 2 diabetes with complications" ) ,
- HIV or AIDS , Tuberculosis (**R.ADM.**CORMBID_CONDITIONS1/2/3/4/OTHER = "Tuberculosis")
- Admission APACHEII score (see Appendix),
- Admission SOFA score (seeAppendixx),
- Previous vaccination (**R_SRASMT.** COVID_VACCINATION_RECEIVED),
- Previous hospitalization this episode (**R_ADM**. READMISSION),
- Duration of symptoms on admission to this hospital (**R_SRASMT.** SYMPTOM_DATE - **R_ADM.**DATE_OF_ADMISSION_HOSPITAL ),
- SpO2 on admission to this hospital **(R_SRASMT**. SATURATION),
- Use of steroids on admission to this hospital (R_SRASMT. CORTICOSTEROIDS)
- Respiratory support at enrolment **(ESC**.BASELINE)

Intervention details will be summarized as follows

- Time from enrolment to first proning session, Time from admission to enrolment, Total number of proning days (I.e. days where proning/visits took place).
- Duration of proning in a 24 hour period [whilst eligible to be proning]

#### Patient acceptability questionnaire

On a scale of 1-10, how did you find lying in the prone position [1 extremely uncomfortable – 10 comfortable]

1 --- 2 --- 3 --- 4 --- 5 --- 6 --- 7 --- 8 --- 9 --- 10

On a scale of 1- 10, how did you find getting into the prone position [1 extremely difficult, needed a lot of help – 10 very easy, could do this unaided]

1 --- 2 --- 3 --- 4 --- 5 --- 6 --- 7 --- 8 --- 9 --- 10

On a scale of 1-10, how did you find comfortable did you find the monitoring equipment [1 extremely uncomfortable – 10 comfortable]

1 --- 2 --- 3 --- 4 --- 5 --- 6 --- 7 --- 8 --- 9 --- 10

On balance, whilst you were receiving oxygen which position do you prefer to lie in for most of the day time

- 1. Prone position
- 2. Supine position
- 3. Side position
- 4. Sitting up
- 5. No preference

On balance, whilst you were receiving oxygen which position do you prefer to lie in for most of the night time

- 1. Prone position
- 2. Supine position
- 3. Side position
- 4. Sitting up
- 5. No preference

Adverse events related to the intervention include

1. Pressure sore
2. Line displacement
3. Severe oxygen desaturation
4. Facial oedema
5. Arrhythmia
6. Hypotension
7. Peripheral nerve injuries
8. Barotrauma
9. Hospital-acquired pneumonia
10. Vomiting

#### Vietnamese guideline of diagnosis and treatment COVID-19, Vietnamese Ministry of Health 06OCT21

##### Clinical severity level

**Moderate:**

Clinical: signs of pneumonia with dyspnoea, respiratory rate 20 - 25 breaths/minute, crackle rales, no signs of severe respiratory failure, $SpO_2$ 94-96% on room air, conscious. Fast or slow pulse rate, tachycardia, normal blood pressure.

**Severe:**

Clinical: signs of pneumonia and accompanied by one of the following: respiratory rate > 25 breaths/minute, severe dyspnoea, accessory respiratory muscle, $SpO_2$ < 94% on room air, tachycardia or bradycardia, normal or high blood pressure, irritable or exhausted, tired.

##### Example of information to be given to patients about positioning

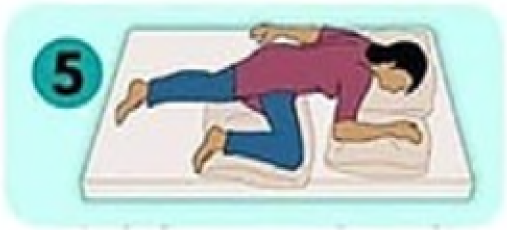

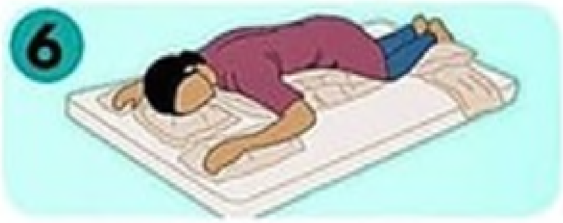
